# Antivenom accessibility impacts mortality and severity of Brazilian snake envenomation: a geospatial information systems analysis

**DOI:** 10.1101/2020.10.13.20211730

**Authors:** Jinny Jing Ye, João Felipe Hermann Costa Scheidt, Luciano de Andrade, Thiago Augusto Hernandes Rocha, Hui Wen Fan, Wuelton Monteiro, Ricardo Palacios, Catherine Ann Staton, João Ricardo Nickenig Vissoci, Charles John Gerardo

## Abstract

**Background:** In 2017, the World Health Organization declared the snakebite envenomation as a neglected tropical disease. Annually, snakebite envenomation causes approximately 400,000 permanent disabilities and 95,000 deaths worldwide. People with the greatest risk of envenomation lack access to adequate health care, including treatment with antivenom. We developed an analysis of accessibility to antivenom in Brazil in order to verify the impacts on mortality.

**Methods and Findings:** Information about number of accidents, deaths, antivenom, medical assistance, and species, were retrieved from the Brazilian Health Informatics Department (DATASUS) from 2010 to 2015 and analyzed using geostatistics to evaluate the association between snakebite accidents and mortality. An Spatial analysis using Global Moran’s I was performed in order to verify the presence of spatiality as an independent variable to the distribution of the accidents. In addition, we also tested three different analysis of regression using Ordinary Least Square (OLS), Spatial Error, and Geographically Weighed Regression (GWR), together with the information obtained from the DATASUS and sociodemographic indicators, to verify the spatial-temporal distribution of envenomation cases and time to reach the healthcare centers. The regression presenting the lowest Akaike Criterion Information (AIC), highest adjusted R^2^, and variables with p < 0.05 was selected to represent our model. Lastly, the accessibility index was performed using 2-step floating catchment area based on the amount of hospital beds and inhabitants. This study revealed 141,039 cases of snakebites, 598 deaths, and mortality rate of 3.13 per 1,000,000 inhabitants. Moreover, GWR presented the best fit (AIC = 55477.56; adjusted R^2^ = 0.55) and showed that illiteracy, income, percentage of urban population, percentage of antivenom, accessibility index for hospital beds with antivenom, proportion of cases with more than 3 hours to reach healthcare are correlated with the mortality rate by snakebite (p < 0.05).

**Conclusion:** This study identified regions affected by snakebite and how the accessibility to antivenom treatment plays an important role in the mortality in Brazil. Public interventions can located to those most vulnerable regions in order to improve the accident outcome.

## Introduction

Snake envenomation is a neglected tropical disease of global importance (1). Estimations around the world are between 1.2 and 5.5 million bites, 421,000 envenomations, 20,000-95,000 deaths, and 400,000 permanent disabilities annually (2). The morbidity and mortality related to this disease primarily impacts resource-limited countries as well as those with lower income earning and other socioeconomic indicators of poverty (3).

In Brazil between 2001 and 2012, 1,192,667 incidents and 2,664 deaths resulting from terrestrial venomous animals (snakes, scorpions, bees, spiders, and caterpillars) were reported. Of these, 28% of all incidences and 54% of deaths were attributed to snake envenomation (4). Of the reported snakebites, 70% of them were from the Bothrops genus. Bothrops envenomation can cause many clinically significant effects from local swelling and bleeding to muscle necrosis, acute kidney injury and cerebral hemorrhage (5) (6). Thus, it becomes important for victims to reach health assistance in time.

Access to snake-specific antivenom is limited because of expensive manufacturing and long transport time to healthcare facility (7). Feitosa et al. 2015 found delays in healthcare access in the Amazon is associated with greater severity and mortality related to snake envenomation (8). Additionally, socioeconomic inequalities have also been linked with lower access health care (9). There is substantial research on the uneven distribution of health care facilities in Brazil (10). However, there is no analysis on accessibility to healthcare and its impacts on snakebite mortality rates in Brazil. Besides, it has not yet been reported how social inequalities are associated with snakebite outcomes.

Therefore, the aim of this study is to analyze the accessibility to antivenom taking into the account the sociodemographics indicators in Brazil in order to verify its impacts on snakebite envenomation mortality using spatial analysis.

## Methods

### Study Design

This study used cross-sectional data on country-wide snakebite envenomations from 2010 to 2015 from National System Identification of Notifiable Diseases (SINAN) stored at DATASUS. We conducted an exploratory spatial analysis to evaluate the geospatial association of snake envenomation mortality in Brazil with socioeconomic predictors (illiteracy, income, percentage of urban area, and human development index), and healthcare access predictors (number of beds, hospitals and health professionals, quantity of antivenom administrated, antivenom coverage, and time to reach the healthcare centers).

### Study population and location

Brazil has a population of 190,732,694 people distributed to 5 regions (North, Northeast, Midwest, Southeast and South) (Figure 1)(11). In recent years, the Brazilian economy has grown rapidly, and the country’s gross domestic product (GDP) is currently the ninth largest globally (9). Brazil has also faced an economic and political crisis since 2010. Despite the growth in the recent years, not all segments of the Brazilian population have benefited uniformly from this rapid development, resulting in substantial income inequality within and between regions of the country, as depicted in Figure 1 (9).

**Figure 1.**
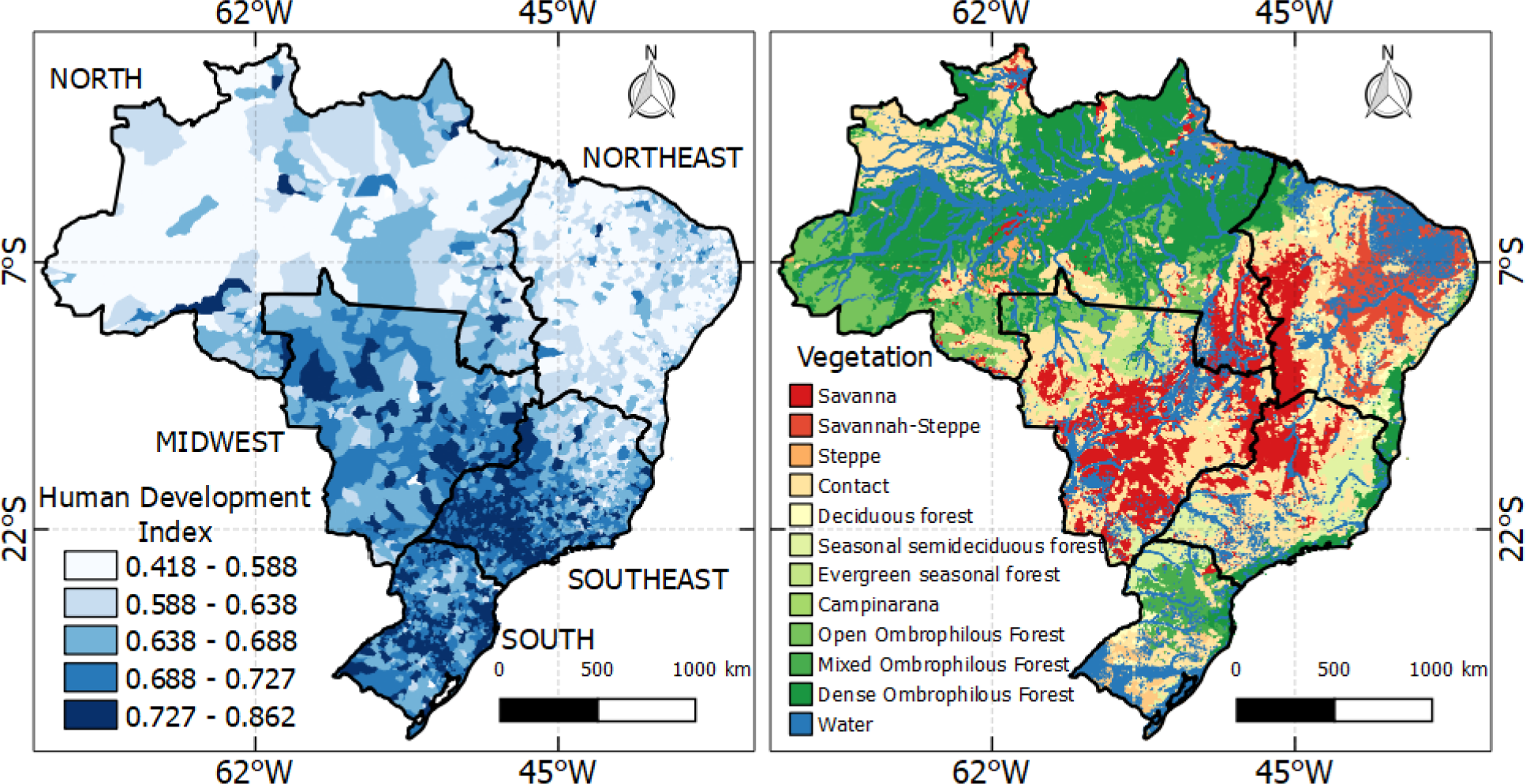
Brazilian gross domestic product on the left and the vegetation on the right.

### Data sources and variables

#### Data extraction

Data regardless sociodemographic indicators were extracted from the Brazilian Institute of Geography and Statistic 2010 Census (11), while information about healthcare centers were obtained from DATASUS and the National Register of Health Facilities (CNES).

#### Snakebite envenomation data

The number of envenomations, hospitalizations from envenomations, deaths from envenomations, time to reach healthcare centers and number of envenomations that were treated with antivenom were collected from SINAN (12). Snakebite outcomes included: mortality, systemic and local complications. An aggregate indicator for snakebite negative outcomes was generated by a sum of the municipality level mortality and complications. Snakebite specific healthcare indicators were: proportion of cases notified more than 3 hours to reach healthcare assistance, and proportions of cases with notified antivenom usage.

#### Data analysis

All analyses in this study were conducted at the municipality-level. Snakebite frequency and complications, socioeconomic indicators and healthcare access were summarized using descriptive statistics.

### Availability of care in hospitals with antivenom

We calculated the availability of hospitals with antivenom by mapping the facilities that received antivenom obtained from the Butantã Institute, agency responsible for producing and distributing antivenom across the country. To evaluate the accessibility to hospitals with antivenom, we used the two-step floating catchment area (2SFCA) method (13). The 2SFCA creates an index of hospitals with antivenom weighted by population in two concurrent steps. In the first step, a buffer was created with a Euclidean 120 kilometer (km) radius surrounding each hospital with antivenom. This buffer was created to collect the population potentially covered in all locations (k) within the threshold. The 120 km range limit was used to estimate a travel distance of two hours to the nearest hospital by car or public transport. The second step consists of summing up the initial ratios in overlapped service areas to measure accessibility for a demand location, where residents have access to multiple supply locations. These steps produced an accessibility index of hospital bed availability with antivenom for each municipality. A higher accessibility index indicates more hospital bed available with antivenom for a given municipality.

### Spatial regression

The Ordinary Least Square (OLS), Spatial Error and geographically weighted regression (GWR) were used to verify the regression with the best fit capable to elucidate the correlation between sociodemographic and healthcare indicators with the mortality rates by snakebite. The regression with the highest adjusted R^2^, lowest AIC, and significant variables (p < 0.05) was chosen as the main model for the study. The spatial autocorrelation and the OLS model were processed using GeoDa software version 1.10.0.8 (Laboratory of Spatial Analysis, University of Illinois at Urbana-Champaign, Urbana, USA). The GWR model was calculated and implemented using R Studio. The maps were generated using QGIS version 2.14.23.

## Results

Between 2010 and 2015, there were a total of 141,039 reported cases of snake envenomation. 598 resulted in death and 27,047 had systemic complications from envenomation. The most frequent cases occurred in the North region (50,102 bites), followed by Southeast (32,474 bites), and least was the South region (12,130 bites). The North region was also in the lead regions with deaths, followed by the Northeast, similar pattern observed to Systemic and Local complications (Table 1).

**Table 1.** Data source and selected variables for the analysis.

| Source | Variables | Website |
| --- | --- | --- |
| **DATASUS**<br>**SINAN** | Snakebite envenomation data:<br>Frequency of cases by municipality;<br>Frequency of hospital admissions by municipalities;<br>Frequency of deaths by municipality;<br>Frequency of local and systemic complications by municipality;<br>Frequency of cases receiving antivenom by municipality;<br>Frequency of cases reaching the hospital with antivenom with time > 3 hours. | http://www2.datasus.gov.br/DATASUS/index.php?area=0203 |
| **CNES**<br>**and**<br>**Butantã Institute** | Availability of care in hospitals with antivenom;<br>Geolocation and hospital beds of all hospitals in the country;<br>List of hospitals receiving antivenom. | http://cnes.datasus.gov.br/ |
| **IBGE** | Socioeconomic indicators:<br>Average income per capita by municipality;<br>Proportion of urban population by municipality;<br>Illiteracy rates by municipality. | https://atlasescolar.ibge.gov.br/ |

DATASUS: Departamento de Informática do Sistema Único de Saúde do Brasil; SINAN: Sistema de Informação de Agravos de Notificação; CNES: Cadastro Nacional de Estabelecimentos de Saúde; IBGE: Instituto Brasileiro de Geografia e Estatística.

According to the registry, the majority of snakebite envenomation received antivenom at some point during their care trajectory. However, the North, Northeast and Midwest had higher proportion of cases with more than 3 hours to reach healthcare (Table 1). Similarly, these were the same regions with lower number of hospitals with lower scores for accessibility for antivenom per population.

**Table 2.** Municipalities indicators of sociodemographics and snakebite envenomations information of each region in Brazil from 2010-2015.

| Indicators | North | Northeast | Midwest | Southeast | South | Amazon |
| --- | --- | --- | --- | --- | --- | --- |
| **Population** | 15.86mi<br>(9.31%) | 53.08mi<br>(27.83%) | 14.05mi<br>(7.37%) | 80.35mi<br>(42.12%) | 27.38mi<br>(14.35%) | 3.48mi<br>(1.82%) |
| **Percentage of Urban areas** | 56.7% | 55.2% | 71.9% | 75% | 60.7% | 83.4% |
| **Envenomation** | 50,102<br>(35.52%) | 31,075<br>(22.03%) | 15,258<br>(10.81%) | 32,474<br>(23.02%) | 12,130<br>(8.60%) | 8,480<br>(6.01%) |
| **Deaths** | 181<br>(30.26%) | 188<br>(31.43%) | 61<br>(10.20%) | 132<br>(22.07%) | 36<br>(6.02%) | 47<br>(7.85%) |
| **Systemic complications** | 6,884<br>(28.62%) | 5,895<br>(24.51%) | 2,721<br>(11.31%) | 6,815<br>(28.34%) | 1,732<br>(7.20%) | 1,603<br>(6.67%) |
| **Proportions of cases with notified antivenom usage** | 47,459<br>(33.7%) | 27,685<br>(19.6%) | 14,490<br>(10.3%) | 29,822<br>(21.2%) | 11,047<br>(7.8%) | 7,893<br>(6.04%) |
| **Proportion of cases notified more than 3 hours to reach healthcare assistance** | 22,809<br>(16.6%) | 9,485<br>(6.9%) | 3,180<br>(2.31%) | 5,143<br>(3.74%) | 1,681<br>(1.22%) | 4,413<br>(10.43%) |
| **Number of hospitals with antivenom** | 231<br>(12.14%) | 416<br>(21.91%) | 189<br>(29.62%) | 564<br>(26.42%) | 502<br>(9.91%) | 40<br>(2.10%) |

**Figure 2.**
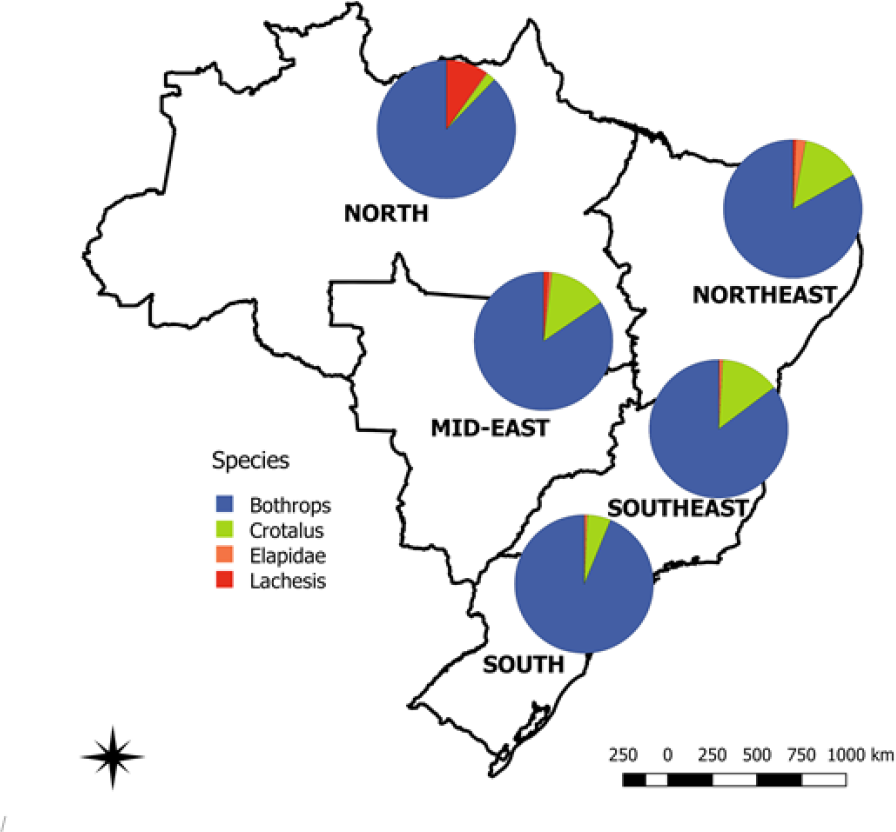
Geographical distribution of snakebite occurrences by snake in Brazil from 2010 to 2015.

**Figure 3.**
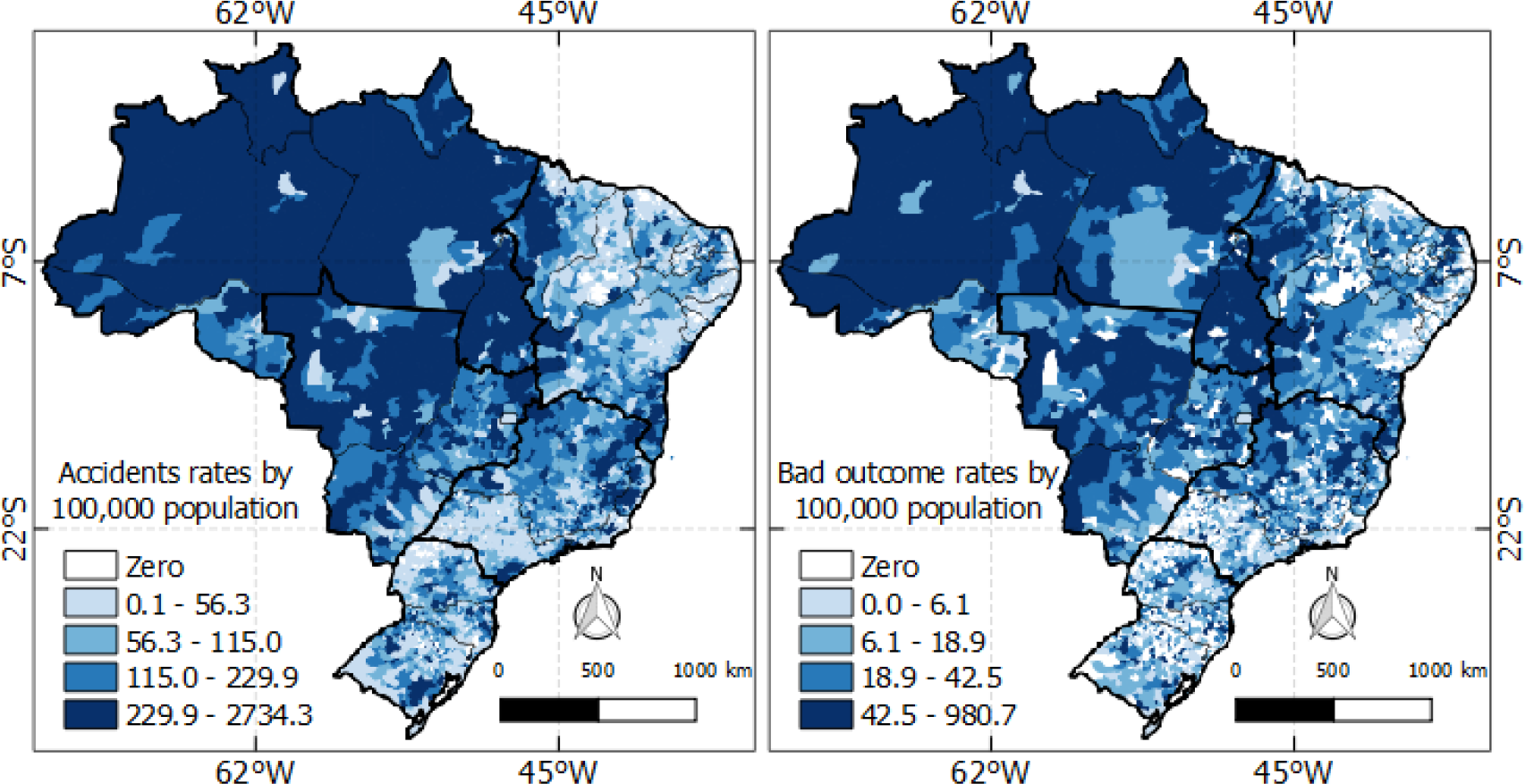
Accidents and deaths per 100,000 inhabitants.

**Figure 4.**
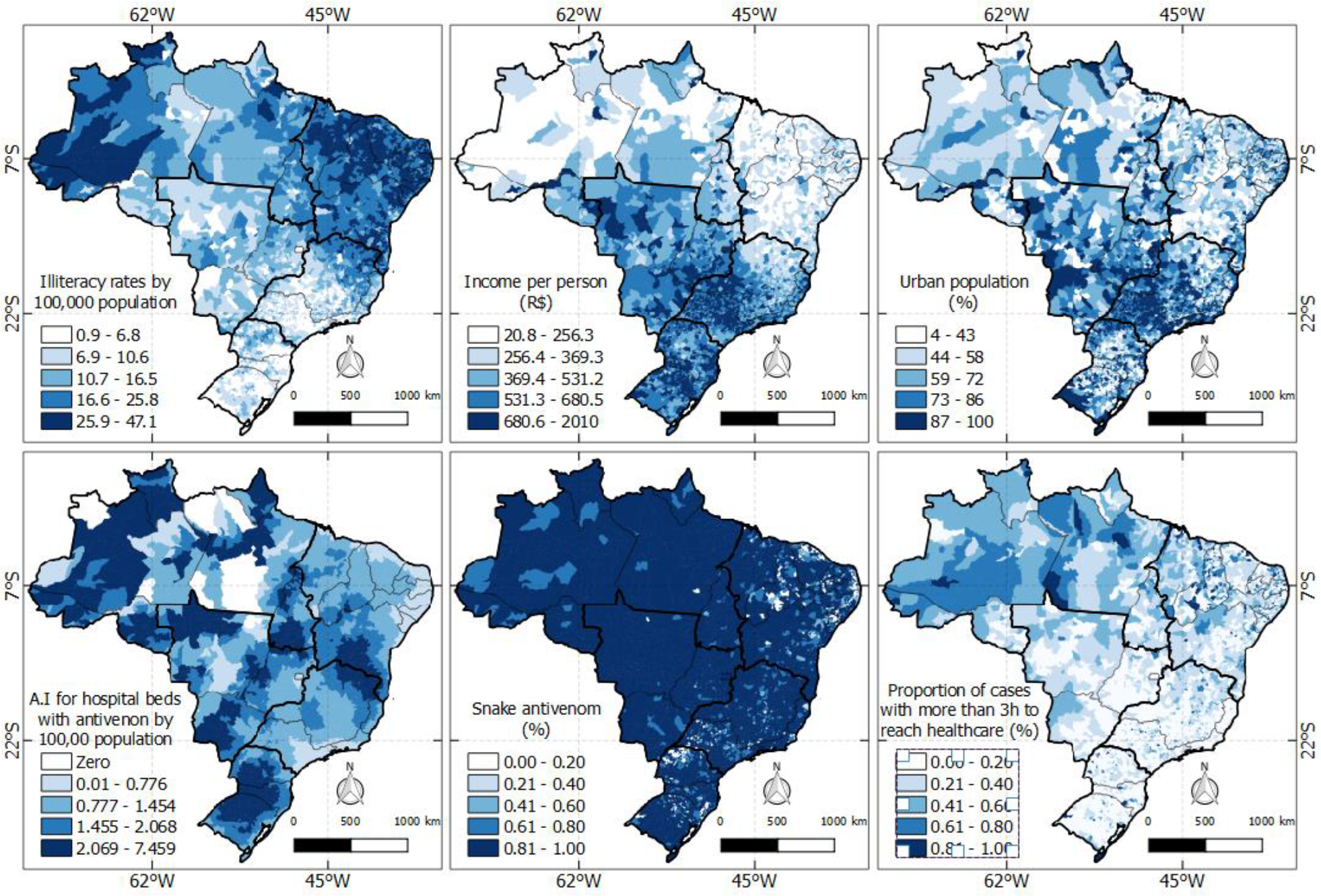
Population-level sociodemographic characteristics and distribution of hospital resources throughout Brazil.

**Figure 5.**
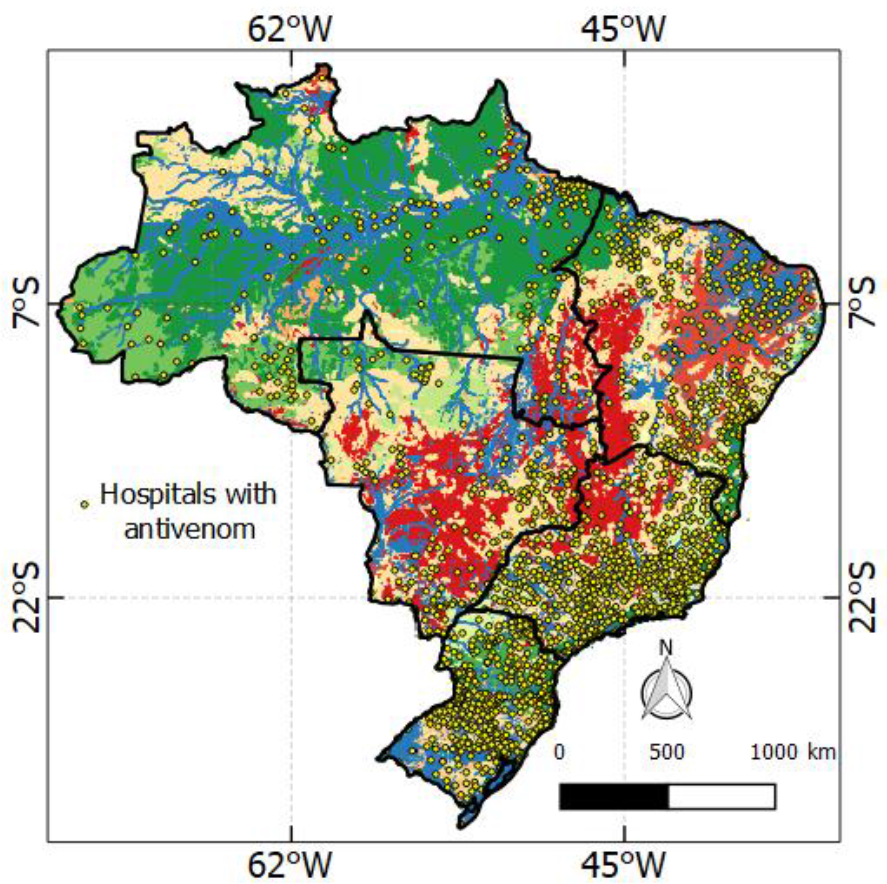
Hospitals with antivenom.

**Table 3.** Comparison between Ordinary Least Square (OLS), Spatial error and Global geographically weighed regression (GWR).

| Variables | OLS |  |  | Spatial Error |  |  | GWR |  |
| --- | --- | --- | --- | --- | --- | --- | --- | --- |
|  | Estimative | P | Standard error | Estimative | p | Standard error | Estimation Mean | Estimation standard deviation |
| Illiteracy | -0.64 | <0.01 | 0.113 | 0.07 | 0.57 | 0.141 | -0.67 | 1.84 |
| Income | -0.05 | <0.01 | 0.004 | -0.02 | <0.01 | 0.004 | -0.04 | 0.06 |
| Percentage of Urban area | -0.24 | <0.01 | 0.035 | -0.37 | <0.01 | 0.034 | -0.23 | 0.32 |
| Antivenom | 26.19 | <0.01 | 2.338 | 13.58 | 2.20 | 0.0 | 26.17 | 24.75 |
| Accessibility index | 4.31 | <0.01 | 0.780 | 1.30 | 1.28 | 0.311 | 4.25 | 21.12 |
| Proportion of cases notified more than 3 hours to reach healthcare assistance | 25.85 | <0.01 | 2.900 | 9.94 | 2.64 | 0.0 | 25.71 | 23.75 |
| AIC | 58546.8 |  |  | 57231.8 |  |  | 55477.56 |  |
| Adjusted R<sup>2</sup> | 0.1127 |  |  | 0.3394 |  |  | 0.5581 |  |

**Figure 6.**
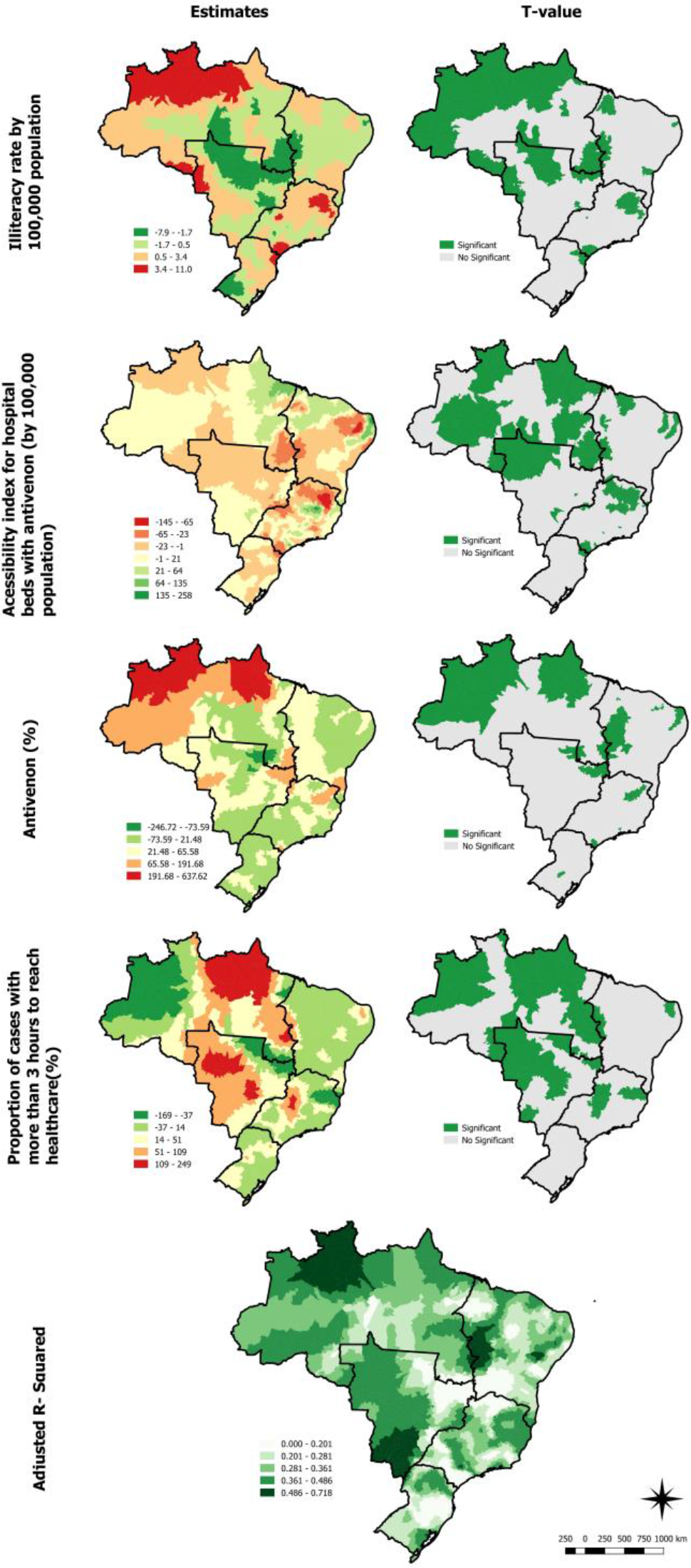
GWR results of significant variables related to the increasing rates of snakebite mortality.

## Discussion

This study is the first nationwide analysis that evaluates the geospatial impact of access to healthcare and antivenom on snakebite mortality. We found a heterogeneous distribution of snakebite mortality and accessibility to healthcare. This is probably attributed to Brazil’s continental dimensions where there are several costumes and cultures (14, 15).

However, the results showed that mortality particularly affected rural and underdeveloped regions of the country. This geographical disparity is corroborated by some studies that revealed that snakebite is a poverty disease and its mortality is correlated with the workforce in agriculture (3, 16). Additionally, the GWR also showed that low illiteracy, income, and percentage of urban area are related to the increased snakebite mortality. Similar study conducted by Suraweera et al (2020) in India also verified that these indicators are correlated with snakebite mortality (17).

Access to antivenom in Brazil varied across the nation with metropolitan areas in the South and Southeast having abundant access compared to the Northwest. Reduced access to antivenom in the North is associated with higher rates of mortality in our study. These are well supplied with antivenom as informed by some studies (18, 19), but the mortality stands out because of the difficulties to reach the healthcare centers, mostly because the main means of locomotion in the amazon is by fluvial transport (20). In addition, those patients that managed to reach healthcare assistance beyond 3 hours were significantly higher in the north suggesting the struggles with the transport.

As for the majority of patients in the country, it was capable to look for health assistance in 3 hours. Our finding is also consistent with findings in Costa Rica where the majority of care received was within 2 hours, even in mountainous and rural regions or other areas with high snakebite incidence. (21)

### Limitations

First, although SINAN is intended to be a nationally comprehensive and accurate database on important notifiable diseases, SINAN is as susceptible as other large population-level databases to reliability and consistency of data collection. Additionally, provider-level reporting of snakebites relies on accurate recognition and diagnosis. Thus, underreporting may not capture the whole scope of this important neglected tropical disease. Second, using a 120km radius to define the catchment area of public hospitals may not be uniform considering rainforest or rural areas cover 60% of Brazil. Despite these limitations, our findings are to our knowledge one of the first studies highlighting the geographical variability of mortality snake envenomations in association with access to antivenom and healthcare.

## Conclusion

In Brazil low illiteracy, income, percentage of urban area, and accessibility to antivenom and healthcare centers are related to the increasing of snakebite mortality. It’s noteworthy to mention that this paper provides regions that lack in those indicators and might useful for the development of better public actions in order to obtain more favorable outcomes.

## Data Availability

No data availability

## References

1. Ranawaka UK, Lalloo DG, de Silva HJ. Neurotoxicity in snakebite--the limits of our knowledge. PLoS Negl Trop Dis. 2013;7(10):e2302.

2. Fox S, Rathuwithana AC, Kasturiratne A, Lalloo DG, de Silva HJ. Underestimation of snakebite mortality by hospital statistics in the Monaragala District of Sri Lanka. Trans R Soc Trop Med Hyg. 2006;100(7):693–5.

3. Harrison RA, Hargreaves A, Wagstaff SC, Faragher B, Lalloo DG. Snake envenoming: a disease of poverty. PLoS Negl Trop Dis. 2009;3(12):e569.

4. Chippaux JP. Epidemiology of envenomations by terrestrial venomous animals in Brazil based on case reporting: from obvious facts to contingencies. J Venom Anim Toxins Incl Trop Dis. 2015;21:13.

5. Cruz LS, Vargas R, Lopes AA. Snakebite envenomation and death in the developing world. Ethn Dis. 2009;19(1 Suppl 1):S1-42-6.

6. de Oliveiraa SS, de Souza Sampaioa V, Sachetta JdAG, Campos E, Fanc HW, de Lacerdaa MVG, et al. Snakebites in the Brazilian Amazon: Current Knowledge and Perspectives. 2016.

7. Chippaux JP. Estimating the global burden of snakebite can help to improve management. PLoS Med. 2008;5(11):e221.

8. Feitosa EL, Sampaio VS, Salinas JL, Queiroz AM, da Silva IM, Gomes AA, et al. Older Age and Time to Medical Assistance Are Associated with Severity and Mortality of Snakebites in the Brazilian Amazon: A Case-Control Study. PLoS One. 2015;10(7):e0132237.

9. Rasella D, Basu S, Hone T, Paes-Sousa R, Ocké-Reis CO, Millett C. Child morbidity and mortality associated with alternative policy responses to the economic crisis in Brazil: A nationwide microsimulation study. PLoS Med. 2018;15(5):e1002570.

10. Garcia-Subirats I, Vargas I, Mogollon-Perez AS, De Paepe P, da Silva MR, Unger JP, et al. Inequities in access to health care in different health systems: a study in municipalities of central Colombia and north-eastern Brazil. Int J Equity Health. 2014;13:10.

11. IBGE. Instituto Brasileiro de Geografia e Estatística. Séries estatísticas & séries históricas. Rio de Janeiro. 2018.

12. Bochner R, Fiszon JT, Machado C. A profile of snake bites in Brazil, 2001 to 2012. 2014.

13. Wang F, Luo W. Assessing spatial and nonspatial factors for healthcare access: towards an integrated approach to defining health professional shortage areas. Health Place. 2005;11(2):131–46.

14. United Nations DoEaSA, Population Division. World Population Prospects: The 2017 Revision. In: United Nations DoEaSA, Population Division, editor. 2017.

15. Bochner R. The international view of envenoming in Brazil: myths and realities. Journal of Venomous Animals and Toxins including Tropical Diseases. 2013;19(1):29.

16. Babo Martins S, Bolon I, Chappuis F, Ray N, Alcoba G, Ochoa C, et al. Snakebite and its impact in rural communities: The need for a One Health approach. PLoS Negl Trop Dis. 2019;13(9):e0007608.

17. Suraweera W, Warrell D, Whitaker R, Menon G, Rodrigues R, Fu SH, et al. Trends in snakebite deaths in India from 2000 to 2019 in a nationally representative mortality study. Elife. 2020;9.

18. Gutiérrez JM, Williams D, Fan HW, Warrell DA. Snakebite envenoming from a global perspective: Towards an integrated approach. Toxicon. 2010;56(7):1223–35.

19. Gutiérrez JM, Fan HW, Silvera CLM, Angulo Y. Stability, distribution and use of antivenoms for snakebite envenomation in Latin America: Report of a workshop. Toxicon. 2009;53(6):625–30.

20. Morgado AV, Portugal LdS, Mello AJR. Acessibilidade na Região Amazônica através do transporte hidroviário. Journal of Transport Literature. 2013;7:97–123.

21. Hansson E, Sasa M, Mattisson K, Robles A, Gutiérrez JM. Using geographical information systems to identify populations in need of improved accessibility to antivenom treatment for snakebite envenoming in Costa Rica. PLoS neglected tropical diseases. 2013;7(1):e2009.

